# Systematic literature review of the use of generative AI in health economic evaluation

**DOI:** 10.1101/2025.04.25.25326412

**Authors:** Steve Sharp, Kusal Lokuge, Jamie Elvidge, Tom Hudson, Dalia Dawoud

## Abstract

**Objectives:** Generative AI (GenAI) has emerged in the current decade as a paradigm-shifting technology with potential to transform the process of health economic evaluation (HEE), a resource-intensive element of health technology assessment. This systematic literature review aims to identify the current applications of GenAI in HEE and its potential advantages, challenges and limitations.

**Methods:** We searched Medline, Embase, EconLit, Cochrane Library, International HTA database and Epistemonikos for English-language, publicly available literature without date restrictions, and hand-searched the ISPOR presentations database (2023 to 2025) for articles describing or investigating the use of GenAI in HEE. Quantitative data on performance outcomes were collected along with qualitative data on stakeholder opinions and experience.

**Results:** We identified 25 eligible studies: 18 primary studies, 6 narrative reviews and 1 expert opinion piece. The primary studies comprised 16 case studies and 2 qualitative studies. Over 90% of studies were conference abstracts published in 2024 from commercial authors. The emphasis across studies was on early exploratory research, particularly model replication. Where reported, execution time (3 studies), accuracy and error rate (7 studies), and user experience (4 studies) showed promising results across multiple use cases but there is a high risk of bias inherent in relying on conference abstracts with limited reporting, which warrants cautious interpretation.

**Conclusion:** The current evidence landscape has revealed potential benefits of GenAI across multiple applications to health economics, but only sparse dissemination of early case study findings via conference submissions. Further research is needed to validate all use cases and to address perceived barriers to implementation.

## 1 Introduction

Developments in the field of artificial intelligence (AI) have accelerated rapidly since the popularisation of generative AI (GenAI) platforms built on large language models (LLMs) in 2022. GenAI’s capability to create new and original content in response to user prompts, by using LLMs that learn from existing data patterns, has transformative potential in the healthcare sector (Moor et al. 2023). Exploration of applying AI in healthcare is already accumulating a large body of research evidence (Alami et al. 2020) and recently focus has shifted to utilising GenAI, including its role in the methods of health technology assessment (HTA).

The National Institute for Health and Care Excellence (NICE) has produced a position statement (NICE 2024a) on the use of AI to set out principles for using AI in evidence generation and reporting of evidence in HTA submissions, reflecting not only the growing interest in AI but also the need for its judicious use. A further statement of intent (NICE 2024b) sets out a flexible framework for future HTA methods development that will adapt to changes in this fast-moving field. Health economic evaluation (HEE) is a resource-intensive and sometimes error-prone element of HTA submissions (Radeva et al. 2020), where GenAI could offer substantial benefits, subject to scrutiny of its own performance compared to the current standard approach. It has the potential to improve efficiency and quality through automation, such as in generating the code for economic models based on text-based prompts (Reason et al. 2024) and, more recently, using model reports (Ayer et al. 2024). It has also demonstrated promise in automating network meta-analysis (Reason et al. 2024), conceptualising economic models, and sourcing input parameters from the literature in a quick and efficient way (Fleurence et al. 2025). Clearly, HEE comprises multiple tasks where GenAI offers potential to improve both efficiency, through quantifiable enhanced performance and time savings, and qualitative user experience, by negating the need for manual completion of traditionally labour-intensive tasks.

We conducted a systematic literature review to establish a baseline of evidence on the different use cases of GenAI in economic evaluations, which will provide a foundation for tracking its evolution and impact over time. As a highly topical subject area with a dynamic evidence base, a living approach (Elliott et al. 2017) is warranted, and we aim to update the review in the near future to capture the accumulating research across the range of study designs and publication types. Mapping the research landscape will provide insights into the more popular use cases of GenAI and highlight priority gaps for further research.

The specific aims of the review are to:

1. Identify and describe the current applications of GenAI in HEE.
2. Identify potential advantages, challenges and limitations associated with the use of GenAI in HEE.

## 2 Methods

Following a pre-specified review protocol, we conducted a systematic search of Medline, Embase, EconLit, Cochrane Library, INAHTA International HTA database and Epistemonikos to identify English-language, publicly available literature without date restrictions. Although LLMs saw significant advancements from 2022, our search was extended to all dates to capture any earlier exploratory studies. The first search was conducted in November 2024 followed by an update search in March 2025.

Citationchaser (Haddaway et al. 2021), which queries the Lens.org database, was used in March 2025 to find references which cited those highlighted as relevant following the initial (November 2024) full-text sift.

We also searched EuropePMC for preprint articles and the PROSPERO database for registered systematic reviews, to reflect the infancy of research into GenAI for economic evaluation.

We browsed the Professional Society for Health Economics and Outcomes Research (ISPOR) conference presentations database for relevant abstracts from conferences held between 2023 and 2024, and manually checked the reference lists of all relevant systematic and non-systematic reviews identified by our search. Reporting was guided by the Preferred Reporting Items for Systematic Review and Meta-analysis (PRISMA) guidelines (Page et al. 2021). Search strategies are summarised in Appendix A.

### Inclusion criteria

We included published studies describing or investigating the use of GenAI, defined as: AI that can create new and original content in response to user prompts or requests by learning from existing data patterns (NICE 2024a, POST). Studies were included if they covered the use of GenAI in economic evaluation in the health science domain, including but not limited to full economic evaluations (including cost-utility, cost-effectiveness, cost-benefit and cost-consequence studies), and cost-comparison studies that include an appropriate demonstration of equal effectiveness. No restrictions were applied to the type or proprietary status of the GenAI tools; publicly available, open-source, and proprietary tools were included. Due to the emergent nature of the evidence base, no restrictions were placed on study design, and conference abstracts, expert opinion pieces and commentaries were included. Systematic and non-systematic reviews were included, where identified.

Studies not describing GenAI in the context of HEE were excluded. Additionally, studies relating to systematic reviewing and sourcing input parameters, including searching, screening and data extraction, were excluded because these broader areas are being extensively studied as separate fields of research.

EPPI Reviewer 5 was used to export, deduplicate and store records from each database. Results were first screened against the selection criteria based on their titles and abstracts by one reviewer. A random 10% selection of titles and abstracts was screened independently by a second reviewer to validate the initial review. All records that potentially met the inclusion criteria were reassessed in full text independently by both reviewers. Discrepancies were resolved by agreement or by input from a third reviewer.

### Data extraction

Data extraction was completed by two reviewers using a pre-specified Microsoft Excel template. We extracted key publication and GenAI characteristics, and data on the main outcomes as reported in the source literature:

- Comparative performance of the GenAI versus manual approaches for modelling elements including but not limited to modelling approach, health states, time horizon, cycle length, incremental cost-effectiveness ratios.
- Time saved relative to manual approaches (mean time saved and reported as mean difference with its 95% CI or as reported in the included studies).
- Difference in errors between the GenAI assisted and manual approaches (percentage difference in errors with 95% CI or as reported in the included studies).
- Qualitative user experience of GenAI versus manual approaches.

### Quality appraisal

Due to the nature of the proposed research question in this review and the wide range of heterogeneous study designs and publication types anticipated, no single risk of bias assessment tool was appropriate to adequately assess bias.

A new quality assessment checklist designed specifically for GenAI tools in economic modelling, the ELEVATE-AI LLMs Framework, was developed during the course of the systematic review by the ISPOR Working Group on Generative AI (Fleurence et al. 2024). This was selected for use with full publications of primary studies, alongside other appropriate appraisal tools for the other study designs, including the SANRA checklist (Baethge et al. 2019), a published and internally validated scale for narrative reviews; the Joanna Briggs Institute checklist for textual evidence-expert opinion (Joanna Briggs Institute 2020); and the CASP qualitative studies checklist (Critical appraisal skills programme 2018).

Applying appropriate quality appraisal checklists aligns with the high standard of reporting recommended in NICE’s position statement on the use of AI in evidence generation (NICE 2024a).

### Synthesis approach

Due to the heterogeneous GenAI tools, use cases, outcomes reported and prompt mechanisms used, data are presented narratively without pooling. The synthesis summarises the number of studies that reported use of GenAI tools; the use case and type of study design; description of the identified tools; and their performance, advantages and limitations. The synthesis of the secondary evidence included analysis of identified reviews to ascertain potential benefits, barriers and challenges to the use of GenAI in HEE. The narrative summary of all the included studies is reported according to the Synthesis Without Meta-analysis (SWiM) guidelines (Campbell et al 2020), where applicable.

## 3 Results

### Included studies

#### Overview of included studies

In total, 2322 studies were eligible for title and abstract review after de-duplication, of which 47 proceeded to full-text screening. These comprised 30 studies from database searching and 17 additional studies identified via conference website and citation searching. Based on full text review, 25 studies were deemed to be eligible for inclusion (Ayer et al. 2024; Boni Z et al. 2024; Chhatwal et al. 2024a; Chhatwal et al. 2024b; Chhatwal et al. 2024c; Dasari et al. 2024, Dolin et al. 2024; Fleurence et al. 2025; Heinz et al. 2024; Knott et al. 2024, Lopez-Bernal et al. 2024; O’Grady et al. 2024; Pandey et al. 2024; Perez-Kempner et al. 2024; Poirrier et al. 2023; Rawlinson W et al. 2024a; Rawlinson W et al. 2024b; Rawlinson W et al. 2024c; Reason T et al. 2024; Srivastava T et al. 2023; Srivastava T et al. 2024a; Srivastava T et al. 2024b; Srivastava T et al. 2024c; Swami et al. 2024; Wu et al. 2024). The selection of evidence is shown in the PRISMA diagram in Appendix B.

Of the 25 included studies, 18 were primary studies, 16 of which were case studies and 2 were qualitative studies. The 6 included reviews were all narrative, with no systematic reviews identified. One opinion piece was also included. Notably, 92% of included studies were conference papers or posters, with just 2 studies published in full (1 case study and 1 narrative review). Almost all studies (23, 92%) were published in 2024. There was also a predominance of industry funded commercial authorship with 24 studies (96%) published by companies, primarily technology consultancies.

#### Primary research

Table 1 below provides the characteristics and results of the included primary studies

**Table 1:**
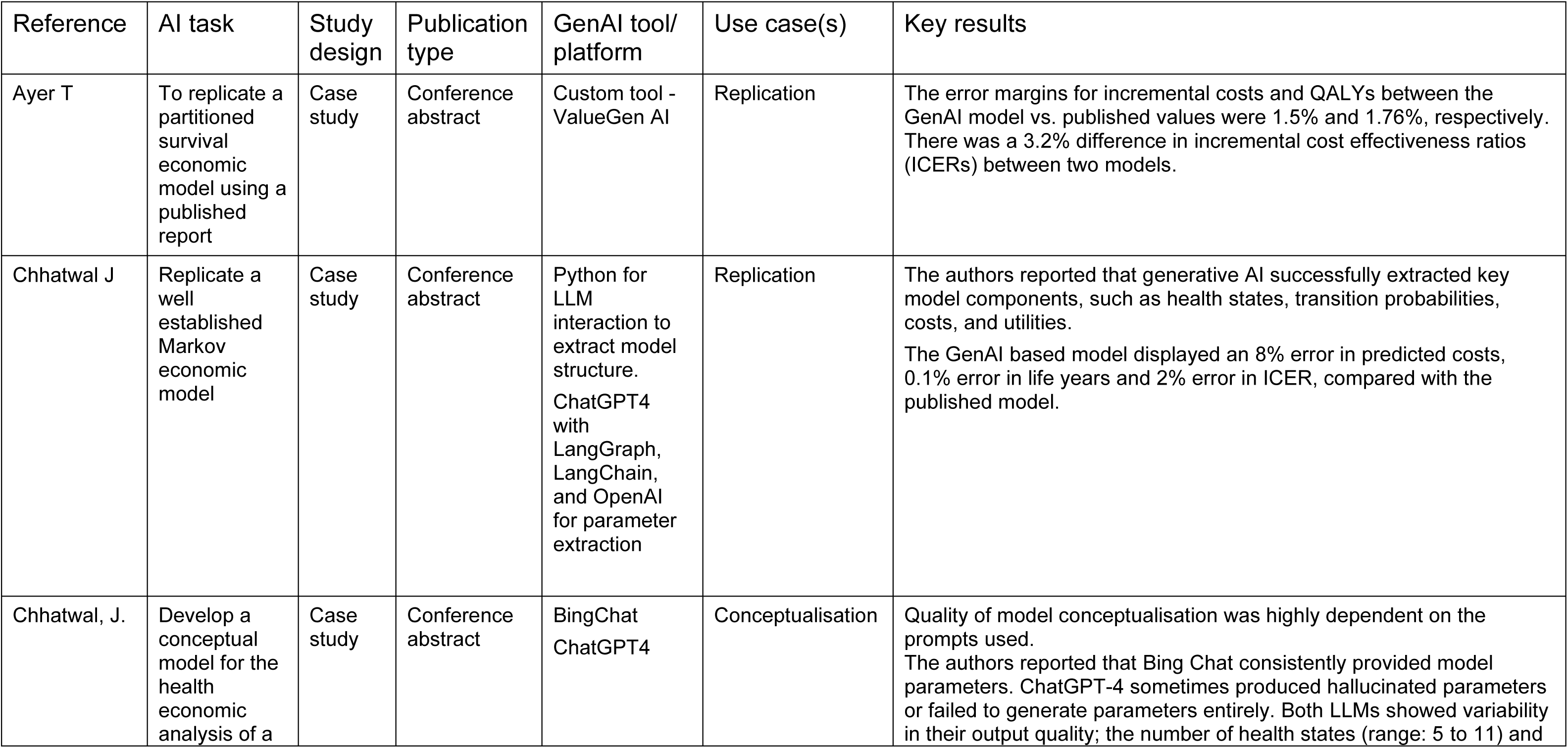
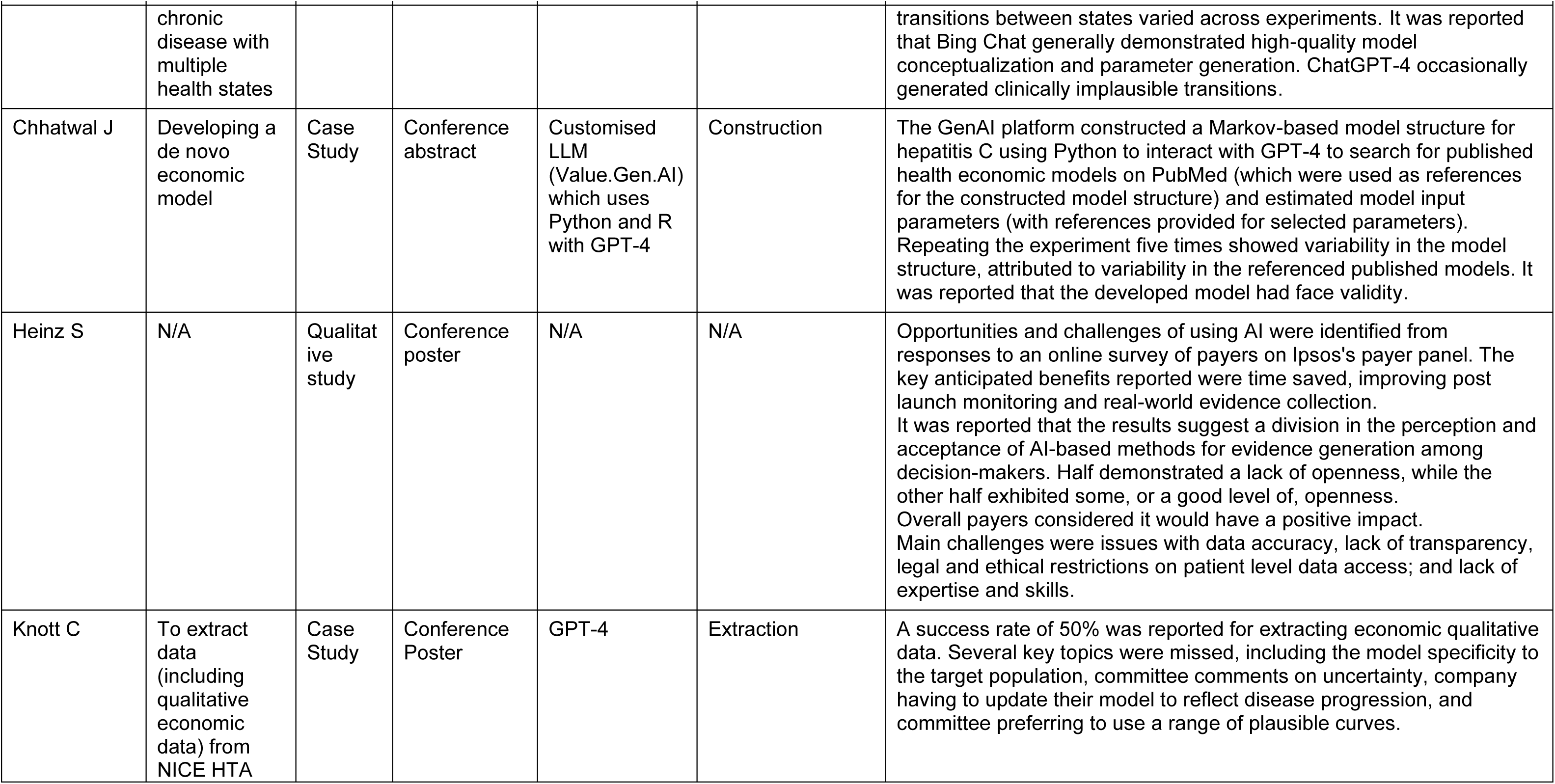
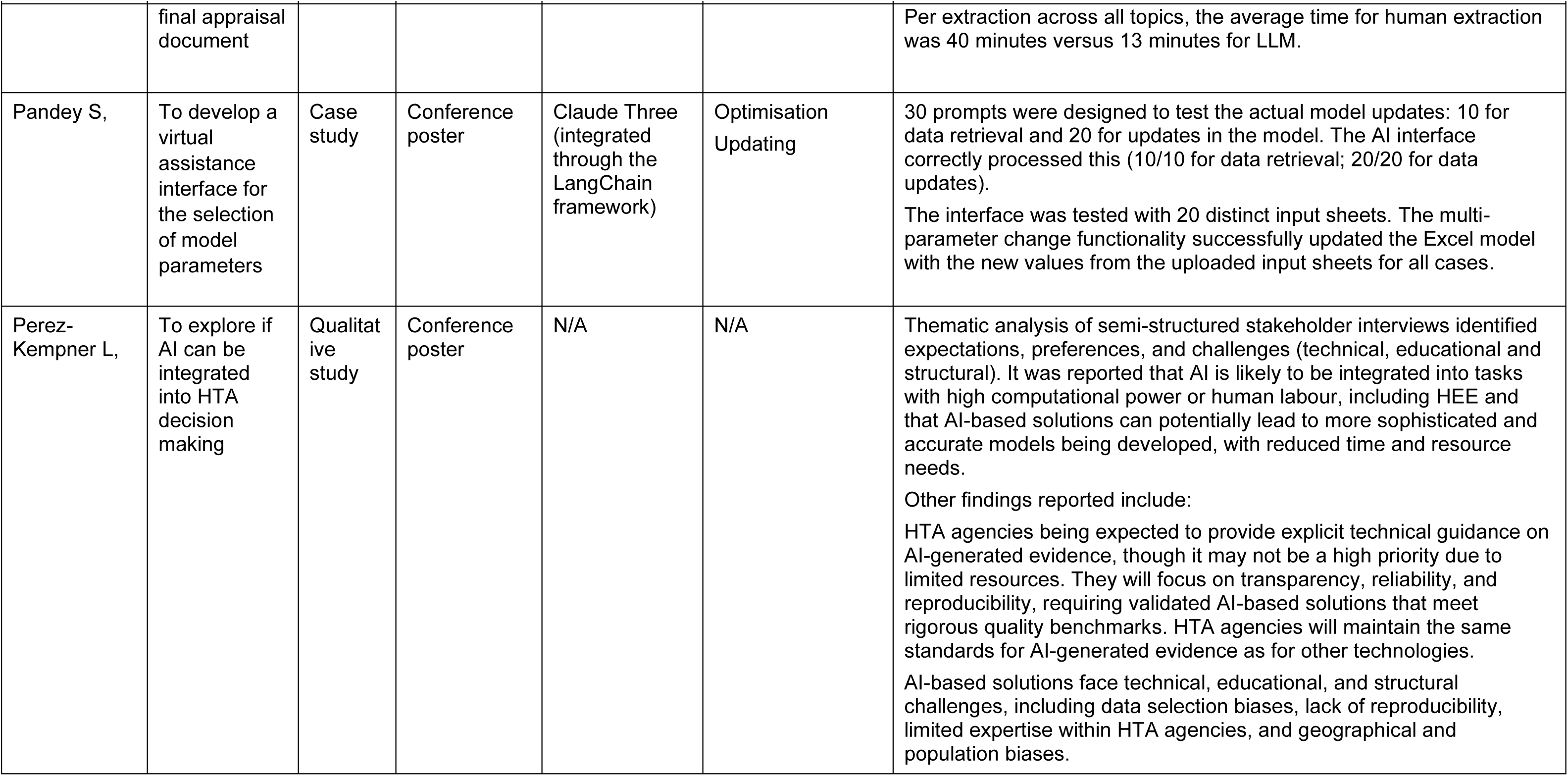
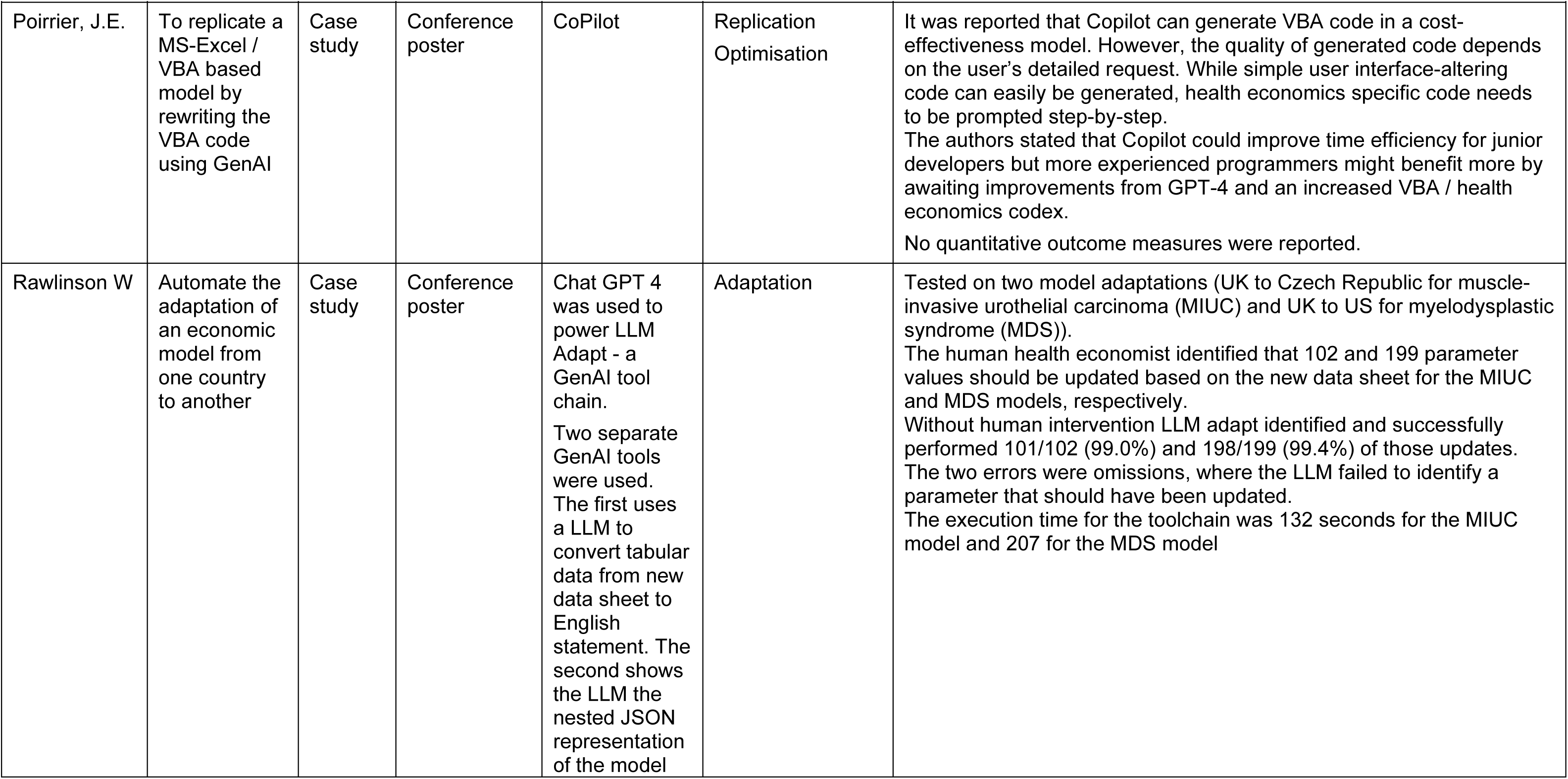
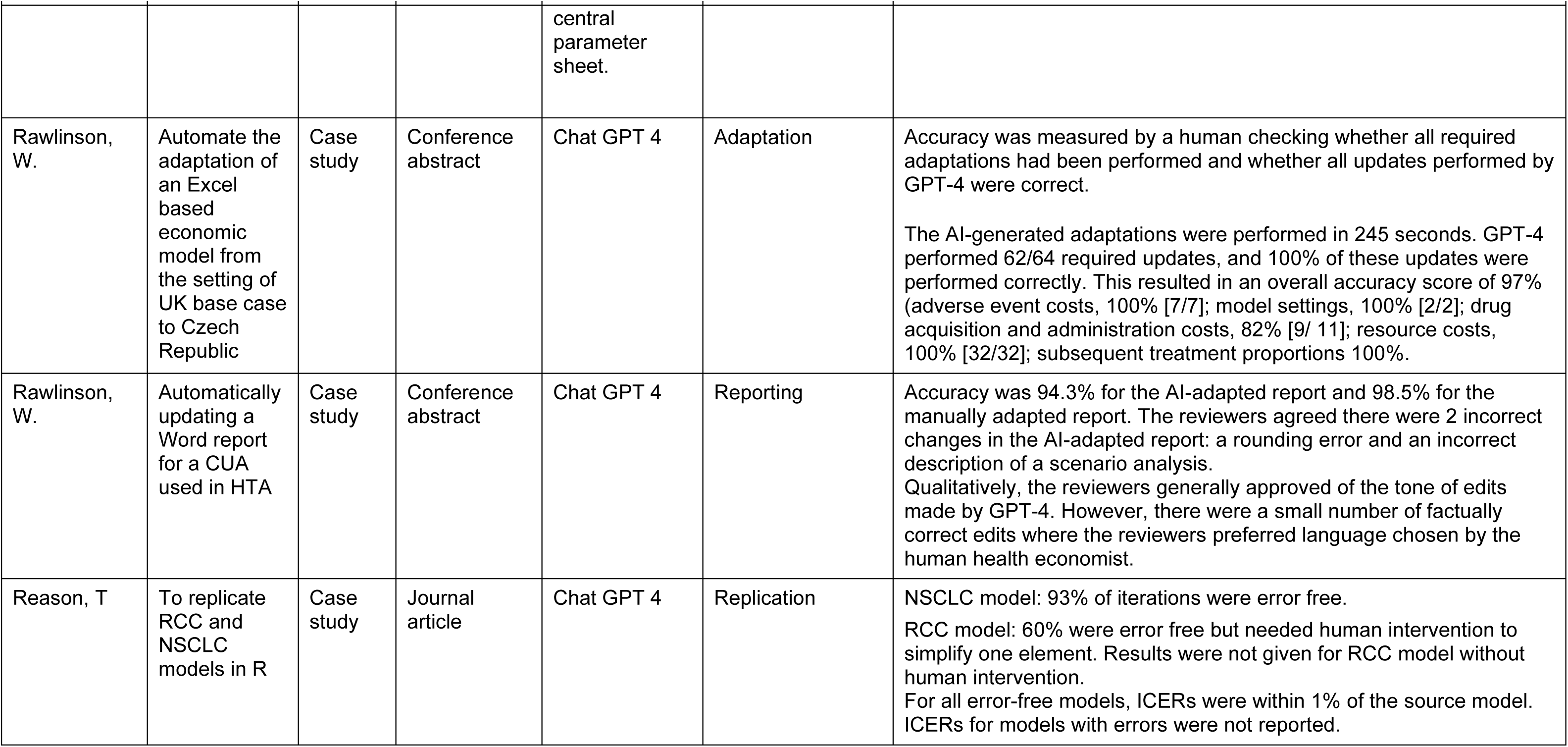
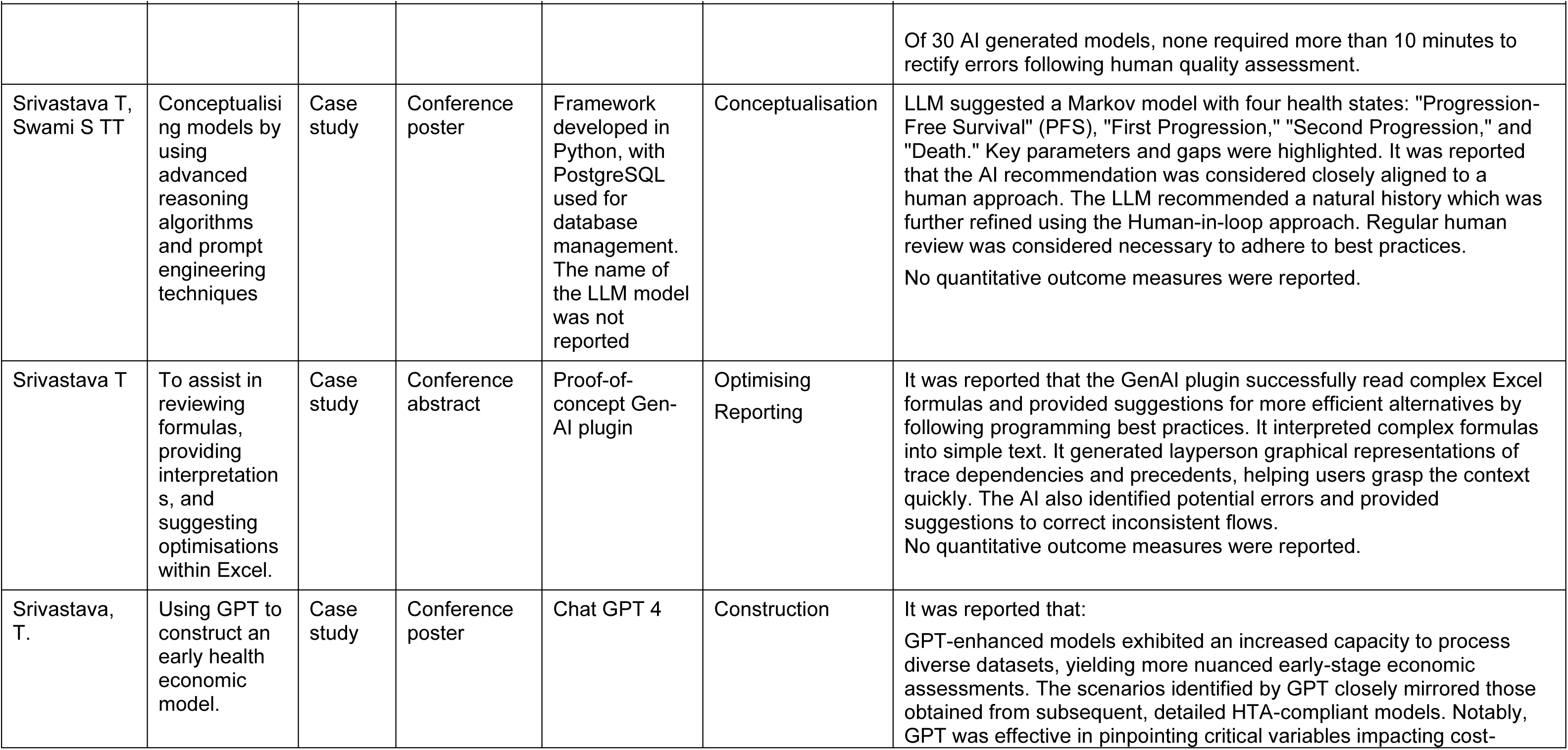
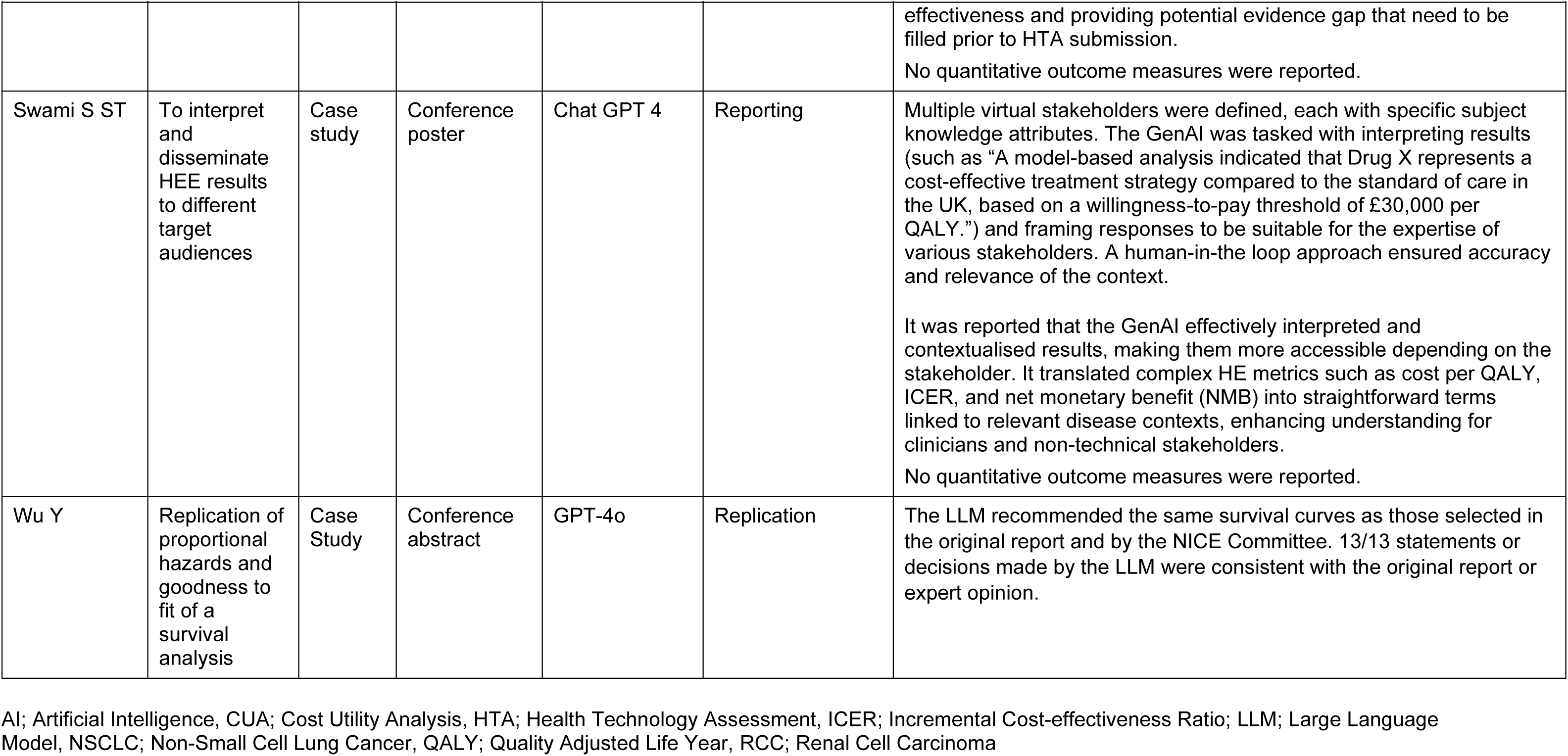
Primary research study characteristics.

| Reference | AI task | Study design | Publication type | GenAI tool/ platform | Use case(s) | Key results |
| --- | --- | --- | --- | --- | --- | --- |
| Ayer T | To replicate a partitioned survival economic model using a published report | Case study | Conference abstract | Custom tool - ValueGen AI | Replication | The error margins for incremental costs and QALYs between the GenAI model vs. published values were 1.5% and 1.76%, respectively. There was a 3.2% difference in incremental cost effectiveness ratios (ICERs) between two models. |
| Chhatwal J | Replicate a well established Markov economic model | Case study | Conference abstract | Python for LLM interaction to extract model structure.<br>ChatGPT4 with LangGraph, LangChain, and OpenAI for parameter extraction | Replication | The authors reported that generative AI successfully extracted key model components, such as health states, transition probabilities, costs, and utilities.<br>The GenAI based model displayed an 8% error in predicted costs, 0.1% error in life years and 2% error in ICER, compared with the published model. |
| Chhatwal, J. | Develop a conceptual model for the health economic analysis of a | Case study | Conference abstract | BingChat<br>ChatGPT4 | Conceptualisation | Quality of model conceptualisation was highly dependent on the prompts used.<br>The authors reported that Bing Chat consistently provided model parameters. ChatGPT-4 sometimes produced hallucinated parameters or failed to generate parameters entirely. Both LLMs showed variability in their output quality; the number of health states (range: 5 to 11) and |

| Reference | AI task | Study design | Publication type | GenAI tool/platform | Use case(s) | Key results |
| --- | --- | --- | --- | --- | --- | --- |
|  | chronic disease with multiple health states |  |  |  |  | transitions between states varied across experiments. It was reported that Bing Chat generally demonstrated high-quality model conceptualization and parameter generation. ChatGPT-4 occasionally generated clinically implausible transitions. |
| Chhatwal J | Developing a de novo economic model | Case Study | Conference abstract | Customised LLM (Value.Gen.AI) which uses Python and R with GPT-4 | Construction | The GenAI platform constructed a Markov-based model structure for hepatitis C using Python to interact with GPT-4 to search for published health economic models on PubMed (which were used as references for the constructed model structure) and estimated model input parameters (with references provided for selected parameters). Repeating the experiment five times showed variability in the model structure, attributed to variability in the referenced published models. It was reported that the developed model had face validity. |
| Heinz S | N/A | Qualitative study | Conference poster | N/A | N/A | Opportunities and challenges of using AI were identified from responses to an online survey of payers on Ipsos's payer panel. The key anticipated benefits reported were time saved, improving post launch monitoring and real-world evidence collection. It was reported that the results suggest a division in the perception and acceptance of AI-based methods for evidence generation among decision-makers. Half demonstrated a lack of openness, while the other half exhibited some, or a good level of, openness. Overall payers considered it would have a positive impact. Main challenges were issues with data accuracy, lack of transparency, legal and ethical restrictions on patient level data access; and lack of expertise and skills. |
| Knott C | To extract data (including qualitative economic data) from NICE HTA | Case Study | Conference Poster | GPT-4 | Extraction | A success rate of 50% was reported for extracting economic qualitative data. Several key topics were missed, including the model specificity to the target population, committee comments on uncertainty, company having to update their model to reflect disease progression, and committee preferring to use a range of plausible curves. |
|  | final appraisal document |  |  |  |  | Per extraction across all topics, the average time for human extraction was 40 minutes versus 13 minutes for LLM. |
| Pandey S, | To develop a virtual assistance interface for the selection of model parameters | Case study | Conference poster | Claude Three (integrated through the LangChain framework) | Optimisation Updating | 30 prompts were designed to test the actual model updates: 10 for data retrieval and 20 for updates in the model. The AI interface correctly processed this (10/10 for data retrieval; 20/20 for data updates).<br>The interface was tested with 20 distinct input sheets. The multi-parameter change functionality successfully updated the Excel model with the new values from the uploaded input sheets for all cases. |
| Perez-Kempner L, | To explore if AI can be integrated into HTA decision making | Qualitative study | Conference poster | N/A | N/A | Thematic analysis of semi-structured stakeholder interviews identified expectations, preferences, and challenges (technical, educational and structural). It was reported that AI is likely to be integrated into tasks with high computational power or human labour, including HEE and that AI-based solutions can potentially lead to more sophisticated and accurate models being developed, with reduced time and resource needs.<br>Other findings reported include:<br>HTA agencies being expected to provide explicit technical guidance on AI-generated evidence, though it may not be a high priority due to limited resources. They will focus on transparency, reliability, and reproducibility, requiring validated AI-based solutions that meet rigorous quality benchmarks. HTA agencies will maintain the same standards for AI-generated evidence as for other technologies.<br>AI-based solutions face technical, educational, and structural challenges, including data selection biases, lack of reproducibility, limited expertise within HTA agencies, and geographical and population biases. |

| Reference | AI task | Study design | Publication type | GenAI tool/platform | Use case(s) | Key results |
| --- | --- | --- | --- | --- | --- | --- |
| Poirrier, J.E. | To replicate a MS-Excel / VBA based model by rewriting the VBA code using GenAI | Case study | Conference poster | CoPilot | Replication<br>Optimisation | It was reported that Copilot can generate VBA code in a cost-effectiveness model. However, the quality of generated code depends on the user's detailed request. While simple user interface-altering code can easily be generated, health economics specific code needs to be prompted step-by-step.<br>The authors stated that Copilot could improve time efficiency for junior developers but more experienced programmers might benefit more by awaiting improvements from GPT-4 and an increased VBA / health economics codex.<br>No quantitative outcome measures were reported. |
| Rawlinson W | Automate the adaptation of an economic model from one country to another | Case study | Conference poster | Chat GPT 4 was used to power LLM Adapt - a GenAI tool chain.<br>Two separate GenAI tools were used. The first uses a LLM to convert tabular data from new data sheet to English statement. The second shows the LLM the nested JSON representation of the model central parameter sheet. | Adaptation | Tested on two model adaptations (UK to Czech Republic for muscle-invasive urothelial carcinoma (MIUC) and UK to US for myelodysplastic syndrome (MDS)).<br>The human health economist identified that 102 and 199 parameter values should be updated based on the new data sheet for the MIUC and MDS models, respectively.<br>Without human intervention LLM adapt identified and successfully performed 101/102 (99.0%) and 198/199 (99.4%) of those updates. The two errors were omissions, where the LLM failed to identify a parameter that should have been updated.<br>The execution time for the toolchain was 132 seconds for the MIUC model and 207 for the MDS model |
| Rawlinson, W. | Automate the adaptation of an Excel based economic model from the setting of UK base case to Czech Republic | Case study | Conference abstract | Chat GPT 4 | Adaptation | Accuracy was measured by a human checking whether all required adaptations had been performed and whether all updates performed by GPT-4 were correct.<br>The AI-generated adaptations were performed in 245 seconds. GPT-4 performed 62/64 required updates, and 100% of these updates were performed correctly. This resulted in an overall accuracy score of 97% (adverse event costs, 100% [7/7]; model settings, 100% [2/2]; drug acquisition and administration costs, 82% [9/ 11]; resource costs, 100% [32/32]; subsequent treatment proportions 100%. |
| Rawlinson, W. | Automatically updating a Word report for a CUA used in HTA | Case study | Conference abstract | Chat GPT 4 | Reporting | Accuracy was 94.3% for the AI-adapted report and 98.5% for the manually adapted report. The reviewers agreed there were 2 incorrect changes in the AI-adapted report: a rounding error and an incorrect description of a scenario analysis.<br>Qualitatively, the reviewers generally approved of the tone of edits made by GPT-4. However, there were a small number of factually correct edits where the reviewers preferred language chosen by the human health economist. |
| Reason, T | To replicate RCC and NSCLC models in R | Case study | Journal article | Chat GPT 4 | Replication | NSCLC model: 93% of iterations were error free.<br>RCC model: 60% were error free but needed human intervention to simplify one element. Results were not given for RCC model without human intervention.<br>For all error-free models, ICERs were within 1% of the source model. ICERs for models with errors were not reported. |

| Reference | AI task | Study design | Publication type | GenAI tool/platform | Use case(s) | Key results |
| --- | --- | --- | --- | --- | --- | --- |
|  |  |  |  |  |  | Of 30 AI generated models, none required more than 10 minutes to rectify errors following human quality assessment. |
| Srivastava T, Swami S TT | Conceptualising models by using advanced reasoning algorithms and prompt engineering techniques | Case study | Conference poster | Framework developed in Python, with PostgreSQL used for database management. The name of the LLM model was not reported | Conceptualisation | LLM suggested a Markov model with four health states: "Progression-Free Survival" (PFS), "First Progression," "Second Progression," and "Death." Key parameters and gaps were highlighted. It was reported that the AI recommendation was considered closely aligned to a human approach. The LLM recommended a natural history which was further refined using the Human-in-loop approach. Regular human review was considered necessary to adhere to best practices.<br>No quantitative outcome measures were reported. |
| Srivastava T | To assist in reviewing formulas, providing interpretations, and suggesting optimisations within Excel. | Case study | Conference abstract | Proof-of-concept GenAI plugin | Optimising Reporting | It was reported that the GenAI plugin successfully read complex Excel formulas and provided suggestions for more efficient alternatives by following programming best practices. It interpreted complex formulas into simple text. It generated layperson graphical representations of trace dependencies and precedents, helping users grasp the context quickly. The AI also identified potential errors and provided suggestions to correct inconsistent flows.<br>No quantitative outcome measures were reported. |
| Srivastava, T. | Using GPT to construct an early health economic model. | Case study | Conference poster | Chat GPT 4 | Construction | It was reported that:<br>GPT-enhanced models exhibited an increased capacity to process diverse datasets, yielding more nuanced early-stage economic assessments. The scenarios identified by GPT closely mirrored those obtained from subsequent, detailed HTA-compliant models. Notably, GPT was effective in pinpointing critical variables impacting cost- effectiveness and providing potential evidence gap that need to be filled prior to HTA submission.<br>No quantitative outcome measures were reported. |
| Swami S ST | To interpret and disseminate HEE results to different target audiences | Case study | Conference poster | Chat GPT 4 | Reporting | Multiple virtual stakeholders were defined, each with specific subject knowledge attributes. The GenAI was tasked with interpreting results (such as “A model-based analysis indicated that Drug X represents a cost-effective treatment strategy compared to the standard of care in the UK, based on a willingness-to-pay threshold of £30,000 per QALY.”) and framing responses to be suitable for the expertise of various stakeholders. A human-in-the loop approach ensured accuracy and relevance of the context.<br>It was reported that the GenAI effectively interpreted and contextualised results, making them more accessible depending on the stakeholder. It translated complex HE metrics such as cost per QALY, ICER, and net monetary benefit (NMB) into straightforward terms linked to relevant disease contexts, enhancing understanding for clinicians and non-technical stakeholders.<br>No quantitative outcome measures were reported. |
| Wu Y | Replication of proportional hazards and goodness to fit of a survival analysis | Case Study | Conference abstract | GPT-4o | Replication | The LLM recommended the same survival curves as those selected in the original report and by the NICE Committee. 13/13 statements or decisions made by the LLM were consistent with the original report or expert opinion. |

AI; Artificial Intelligence, CUA; Cost Utility Analysis, HTA; Health Technology Assessment, ICER; Incremental Cost-effectiveness Ratio; LLM; Large Language Model, NSCLC; Non-Small Cell Lung Cancer, QALY; Quality Adjusted Life Year, RCC; Renal Cell Carcinoma

#### Quality appraisal

The inadequate level of reporting inherent in conference abstracts and the absence of full publications precluded the use of the ELEVATE-AI LLM checklist for all except one of the included case studies. The single full publication (Reason et al. 2024) was appraised as part of the ELEVATE-AI LLM validation (Fleurence et al. 2024) and scored 24/30 across all the domains.

#### Qualitative research on potential GenAI applications

Two qualitative studies published as conference abstracts were identified (Heinz et al. 2024 and Perez-Kempner et al 2024). Heinz et al. (2024) examined the opportunities and challenges of AI as seen by an international payer panel, with results showing a division in the perception and acceptance of AI-based methods for evidence generation in HTA. The key challenges highlighted concerns about lack of transparency in AI algorithms, questions around data reliability, validity, and accuracy, and the absence of guiding regulatory frameworks for AI use. However, the payers anticipated that incorporation of AI in their processes would have a positive impact overall.

Perez-Kempner et al. (2024) interviewed eight stakeholders from different technical backgrounds, including health economists, who concluded that AI could improve efficiency in HTA more broadly, but highlighted the need for HTA agencies to focus on the transparency, reliability and reproducibility of AI-generated evidence. Hence, cross-functional collaboration between AI experts and HTA agencies will be essential in understanding and incorporating AI technology. Several perceived technical, educational, and structural challenges were highlighted, including data selection biases, lack of reproducibility, increased energy consumption, limited expertise within HTA agencies, and geographical and population biases. The current lack of technical guidance from HTA agencies was viewed as both a barrier and an expectation, with internationally referenced HTA agencies expected to shape future policy.

#### Case study synthesis

##### Conceptualisation

Two studies (Chhatwal et al. 2024a, Srivastava et al. 2024a) used LLMs to conceptualise an economic model, with both studies using different prompt mechanisms as explained in Table 1.

Chhatwal et al. (2024a) used BingChat and ChatGPT-4 to conceptualise an economic model, reporting that BingChat demonstrated higher quality model conceptualisation and parameter generation when compared to ChatGPT-4. Srivastava et al. (2024a) used retrieval-augmented generation (RAG) that combined LLM information retrieval (with the RAG database populated with HEOR related guidelines and disease specific documents) with a human-in-the-loop approach. There was insufficient information reported by Chhatwal et al. (2024a) to deduce the accuracy of the conceptualised model when compared to a human approach. However, the quality of model conceptualisation was highly dependent on the prompts used, highlighting the need for expert guidance.

Similarly, Srivastava et al. (2024a) reported qualitatively that the LLM results aligned closely with an expected output from human experts but highlighted the need for human review of the LLM approach to adhere to best practice.

##### Replication

Five studies, comprising 4 conference abstracts (Ayer et al. 2024, Chhatwal et al. 2024b, Poirrier et al. 2023, Wu et al. 2024) and one journal publication (Reason et al. 2024), used LLMs to replicate an existing economic model (one study replicated two economic models, and another replicated a survival analysis of a partitioned survival model). The replication of models can be seen as a preliminary use case, as it potentially validates the use of LLMs to develop, adapt and update economic models. Prompting mechanisms varied between studies as reported in Table 1. Three studies (Reason et al. 2024, Poirrier et al. 2024 and Wu et al. 2024) used publicly available LLMs for model replication, while 2 studies (Ayer et al. 2024 and Chhatwal et al. 2024) used a customised LLM, ValueGen.AI, a GPT-4-based platform using a multi agent pipeline, where data was extracted from a published document.

Error percentages (defined as differences in results between the original manually developed model, and the model replicated by the LLM) were reported in 3 studies and were largely below 10%. An exception to this was the replication of the renal cell carcinoma (RCC) model by Reason et al. (2024) who compared the R scripts of the replicated health economic model with the human developed model to calculate error percentages. Here, despite human intervention to simplify one element, only 60% of iterations were completely error free (87% were error free or with single minor error). However, in both models tested, error-free scripts replicated published incremental cost effectiveness ratios (ICERs) to within 1% of original values. Time to rectify errors was only reported by Reason et al. (2024), with all errors rectified within 10 minutes of quality control detection.

A study (Wu et al. 2024) on replicating a survival analysis in a partitioned survival model did not report error percentages but noted that all decisions made by the LLM relating to assessing proportional hazards and goodness-of-fit were consistent with the original report or expert opinion. However, the authors also recommended further research to generalise these results to data sets with varying levels of maturity and other GenAI models.

One study (Poirrier et al. 2023) did not report quantitative outcomes, acknowledging the need for a quantitative evaluation of the tool before wider adoption by health economists. It also highlighted that issues in code explainability, ownership and licensing need to be clarified.

Two of the five studies (Poirrier et al. 2023 and Reason et al. 2024) highlighted the need for human intervention when replicating an economic model, specifically for probabilistic sensitivity analysis and simplifying model design.

##### Adaptation

Two case studies used LLMs to adapt models from one geographical setting to another using ChatGPT-4 (Rawlinson et al. 2024a, Rawlinson et al. 2024b). Prompting mechanisms differed between the studies as outline in Table 1. Both the studies reported high levels of accuracy, with the 97% to 100% of required parameters adapted correctly (with parameters classified into separate categories such as adverse event costs). The only deviation from this was the adapting of drug and acquisition costs in Rawlinson et al. (2024b) where only 82% of parameters were adapted correctly. Notably, human intervention was not necessary to achieve these levels of adaptation accuracy. The processing time needed to perform the necessary adaptations was extremely low, thereby signalling efficiency gains if GenAI is used to adapt economic models.

##### Construction

Two conference abstracts tested the construction of an early-stage economic model using an LLM. Srivastava et al. (2024b) used Chat GPT-4 while Chhatwal et al. (2024c) used a customised LLM, ValueGen.AI, built using R, Python and Chat GPT-4. There was a lack of clarity on the prompt mechanism used in both studies.

There was also a lack of clarity on the extent to which the model was constructed in Srivastava et al. (2024b), with the LLM reportedly used to identify scenarios to be considered in a cost-effectiveness analysis, key variables impacting cost-effectiveness and evidence gaps. While study accuracy was not quantitatively reported, the abstract did report that the scenarios identified by the LLM mirrored those obtained from subsequent, detailed HTA-compliant model reports.

Chhatwal et al. (2024c) constructed a Markov-based model structure for hepatitis C using published models used as references and estimated model input parameters with references for selected parameters. Repeating the experiment five times led to differences in resulting model structures. The authors cited variations in published models as the main cause of this. It was reported that the model had face validity, although it was unclear as to how this was established.

##### Validation

There were no studies exploring the validation of economic models, either through internal quality assessment and error-checking or external validation relative to clinical data or knowledge, highlighting a potential gap in this area. This aligns with the broader finding in previous reviews that validation is a missing element in many human-led modelling studies (Silva-Illanes and Espinoza. 2018; Altunkaya et al. 2021).

##### Updating

Pandey et al. (2024) developed a virtual interface which used the capabilities of Claude 3, to help model adaptation using a structured prompt mechanism. The developed interface correctly processed 10 data retrieval and 20 model updates with 100% accuracy, highlighting the benefits of designing such a user interface with LLM capabilities to update models.

##### Optimisation

One case study (Srivastava et al. 2024c) investigated how a GenAI based plugin for an Excel model can assist in reviewing formulae, providing interpretations, and suggesting optimisations within Excel. There was a lack of clarity on which LLM was used to build the plugin. No quantitative outcome measures were reported to judge the accuracy or success of the optimisation. However, the abstract reported that the plugin read complex Excel formulae and provided suggestions for more efficient alternatives. The plugin also identified potential errors and provided suggestions to correct inconsistent flows.

The virtual assistance interface (Pandey et al. 2024) used for updating also reportedly served the use case of optimising Excel-based models, with the interface enabling reliable performance of all calculations consistently within a user-friendly environment suitable for non-modellers. However, the outcome measures for user experience were not reported.

##### Reporting and dissemination

Three conference abstracts explored the use of LLMs (Chat GPT4) to report and disseminate cost-effectiveness results (Rawlinson et al. 2024c, Srivastava et al. 2024b, Swami et al. 2024). The studies differed in approach, with Rawlinson et al (2024c) using a LLM to automatically update a Word document report after making changes to the cost utility analysis (CUA), and Swami et al. (2024) using a LLM to disseminate health economic modelling results to different audiences via a paragraph of text. The prompt mechanisms used in all the studies were unclear. The GenAI plugin (Srivastava et al. 2024b) used for optimisation also generated graphical representations of trace dependencies and precedents in the model, which were reported as helping to convey the context to lay users quickly. However, no quantitative outcomes were reported.

Results from Rawlinson et al. (2024c) showed a high level of accuracy for the AI generated report, with it capturing a large number of updates applied to the CUA. Swami et al. (2024) did not quantify the accuracy of the dissemination, but stated that the exercise shows promise, subject to further refinement.

##### Extraction

One study (Knott et al. 2024) looked at the use of GenAI (GPT-4) in the market access analysis task of extracting data from a NICE HTA final appraisal document, to identify trends in decision making and potentially inform future submissions. This included the extraction of quantitative economic data which had a success rate of 100%, and qualitative economic data, which had the lowest success rate (50%) with the extraction failing to capture several key topics as detailed in Table 1. While the success rate for the extraction of qualitative economic data was low, it is worth noting that the success rate of the extraction of other qualitative data (interventions, severity modifier, differentiators, clinical data and recommendations) was much higher, ranging from 92% to 100%. The reasons for these differences were not reported in the study.

#### Secondary research

A total of 6 non-systematic narrative reviews were identified in the literature search, defined as reviews summarising the literature without explicitly following the systematic methods described in the Cochrane handbook.

All the reviews were published in 2024, highlighting the growing research momentum in this new era of GenAI technology. Five of the reviews were published as conference papers. Table 2 details the review characteristics.

**Table 2.**
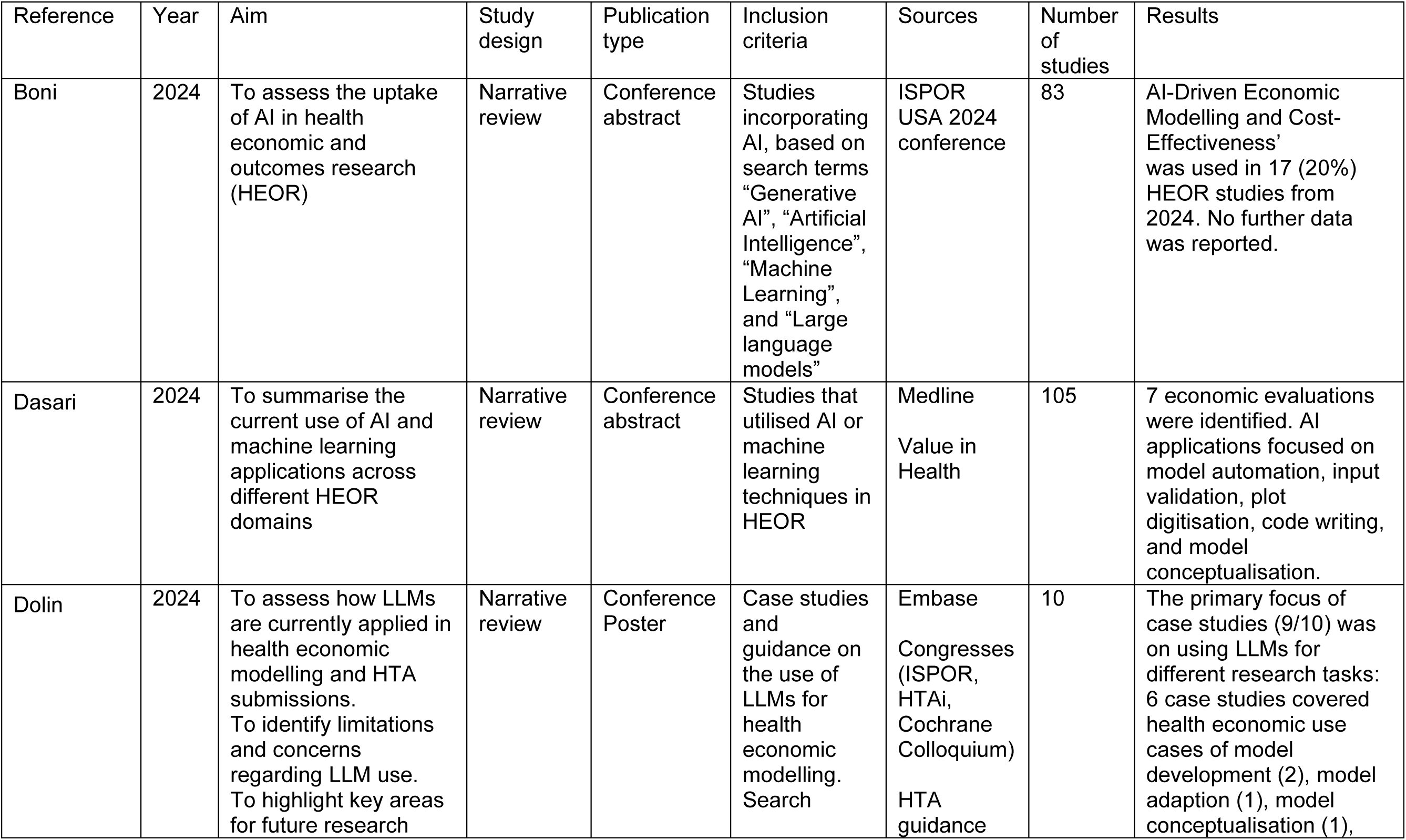
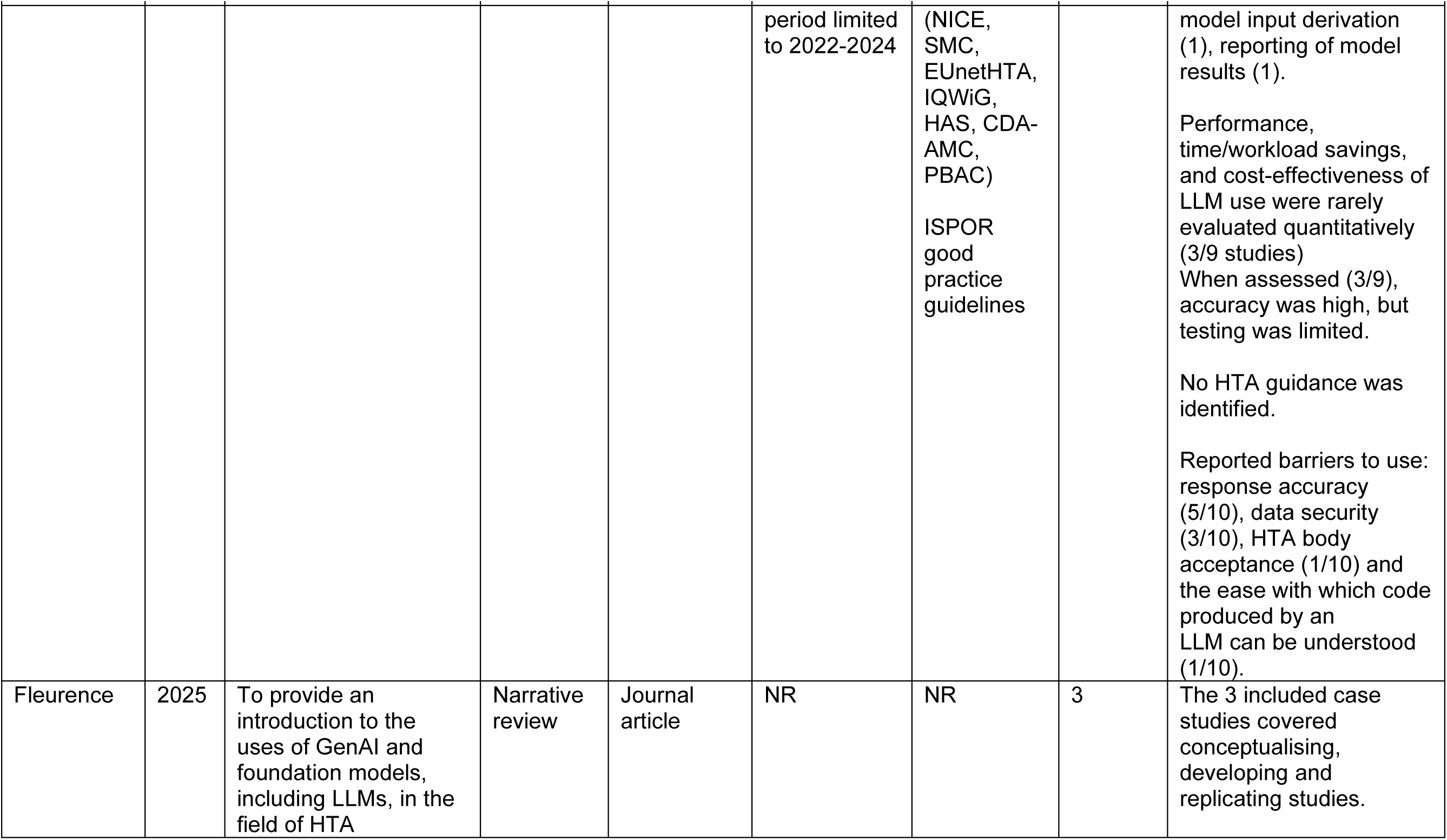
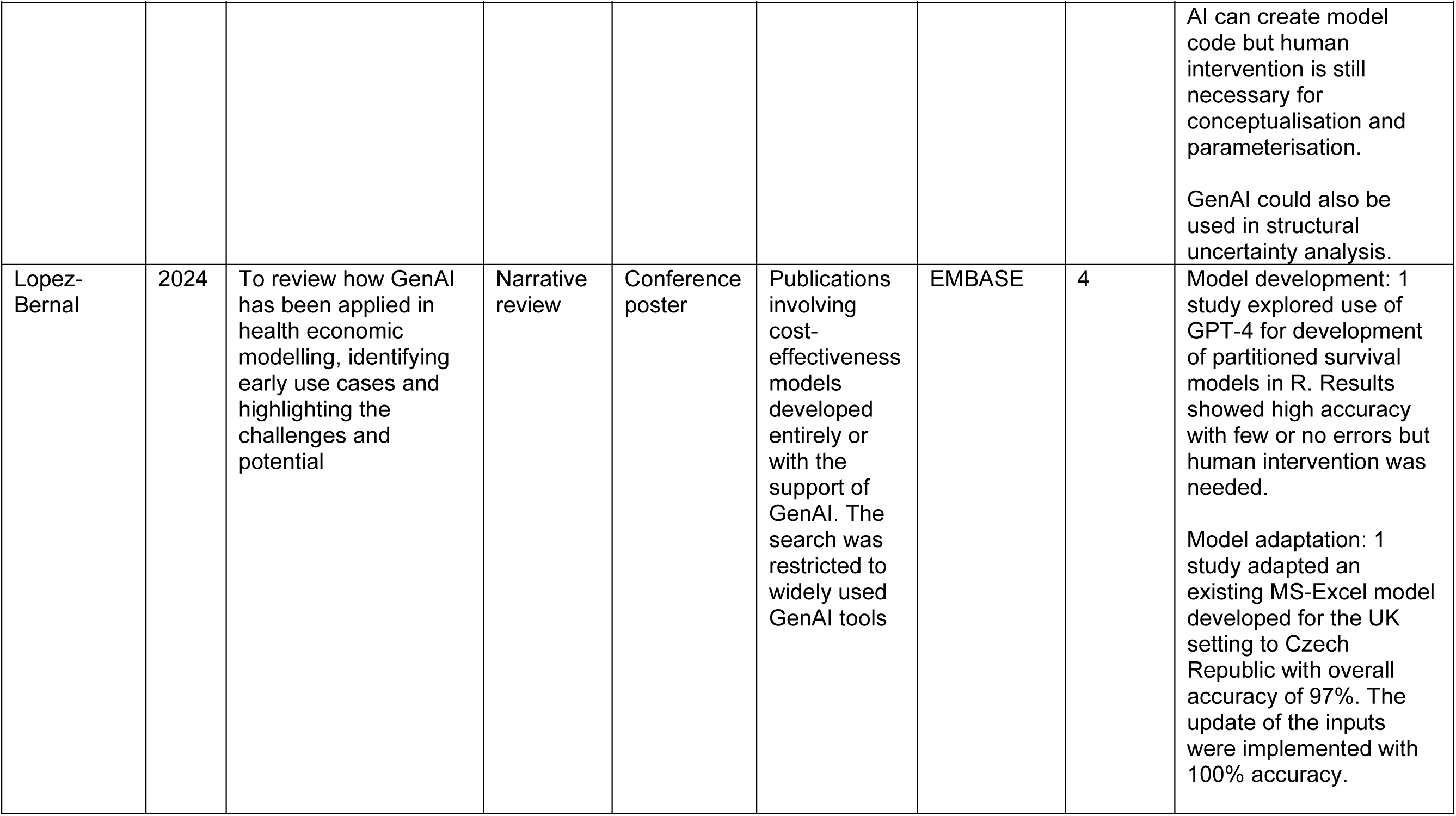
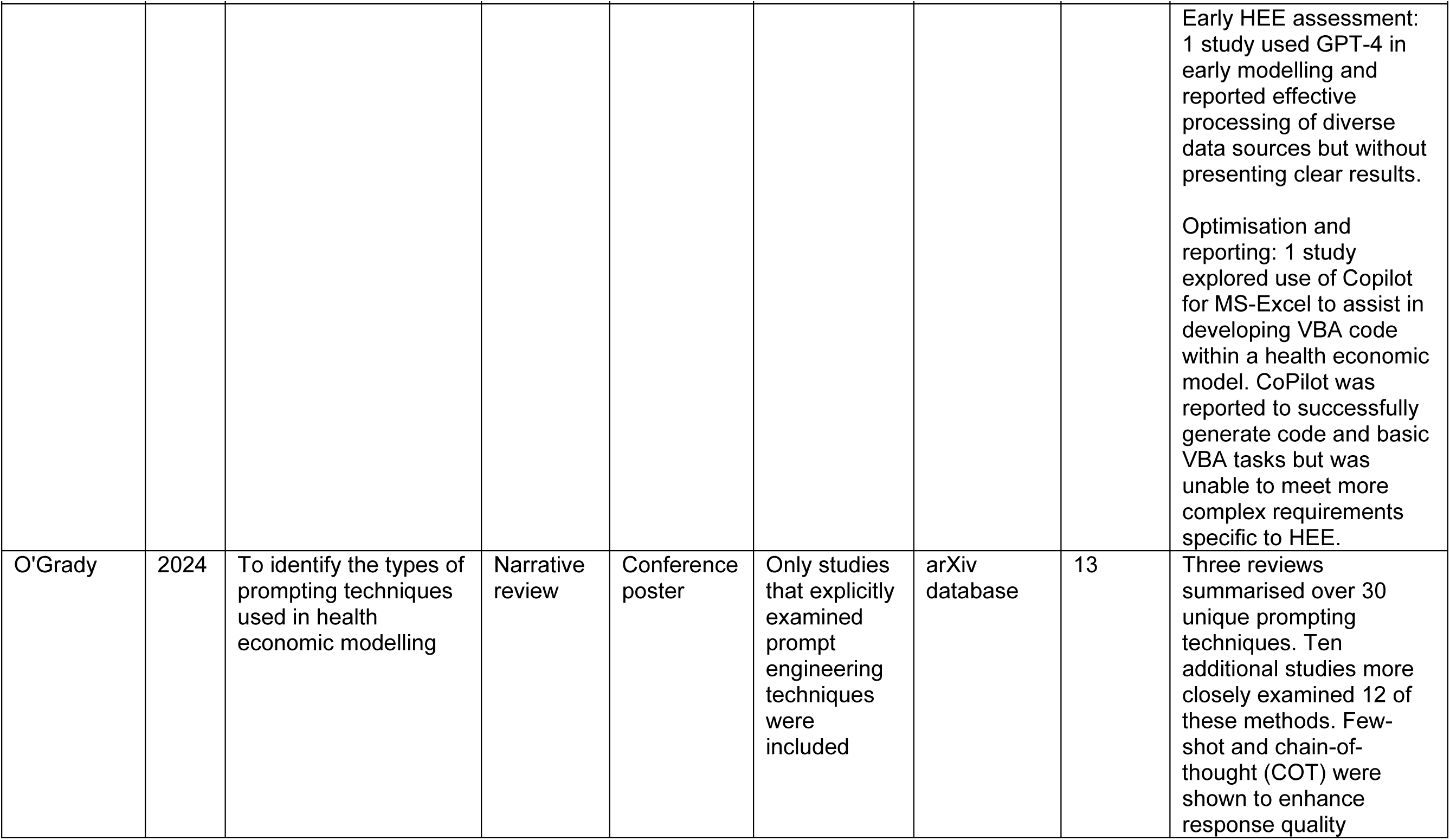
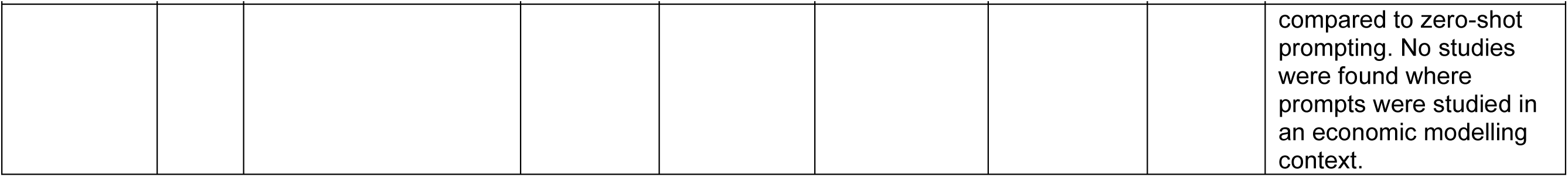
Study Characteristics of included reviews.

| Reference | Year | Aim | Study design | Publication type | Inclusion criteria | Sources | Number of studies | Results |
| --- | --- | --- | --- | --- | --- | --- | --- | --- |
| Boni | 2024 | To assess the uptake of AI in health economic and outcomes research (HEOR) | Narrative review | Conference abstract | Studies incorporating AI, based on search terms “Generative AI”, “Artificial Intelligence”, “Machine Learning”, and “Large language models” | ISPOR USA 2024 conference | 83 | AI-Driven Economic Modelling and Cost-Effectiveness’ was used in 17 (20%) HEOR studies from 2024. No further data was reported. |
| Dasari | 2024 | To summarise the current use of AI and machine learning applications across different HEOR domains | Narrative review | Conference abstract | Studies that utilised AI or machine learning techniques in HEOR | Medline<br>Value in Health | 105 | 7 economic evaluations were identified. AI applications focused on model automation, input validation, plot digitisation, code writing, and model conceptualisation. |
| Dolin | 2024 | To assess how LLMs are currently applied in health economic modelling and HTA submissions.<br>To identify limitations and concerns regarding LLM use.<br>To highlight key areas for future research | Narrative review | Conference Poster | Case studies and guidance on the use of LLMs for health economic modelling. Search | Embase<br>Congresses (ISPOR, HTAi, Cochrane Colloquium)<br>HTA guidance | 10 | The primary focus of case studies (9/10) was on using LLMs for different research tasks: 6 case studies covered health economic use cases of model development (2), model adaption (1), model conceptualisation (1), |
|  |  |  |  |  | period limited to 2022-2024 | (NICE, SMC, EUnetHTA, IQWiG, HAS, CDA-AMC, PBAC)<br>ISPOR good practice guidelines |  | model input derivation (1), reporting of model results (1).<br>Performance, time/workload savings, and cost-effectiveness of LLM use were rarely evaluated quantitatively (3/9 studies)<br>When assessed (3/9), accuracy was high, but testing was limited.<br>No HTA guidance was identified.<br>Reported barriers to use: response accuracy (5/10), data security (3/10), HTA body acceptance (1/10) and the ease with which code produced by an LLM can be understood (1/10). |
| Fleurence | 2025 | To provide an introduction to the uses of GenAI and foundation models, including LLMs, in the field of HTA | Narrative review | Journal article | NR | NR | 3 | The 3 included case studies covered conceptualising, developing and replicating studies. |
|  |  |  |  |  |  |  |  | AI can create model code but human intervention is still necessary for conceptualisation and parameterisation.<br>GenAI could also be used in structural uncertainty analysis. |
| Lopez-Bernal | 2024 | To review how GenAI has been applied in health economic modelling, identifying early use cases and highlighting the challenges and potential | Narrative review | Conference poster | Publications involving cost-effectiveness models developed entirely or with the support of GenAI. The search was restricted to widely used GenAI tools | EMBASE | 4 | Model development: 1 study explored use of GPT-4 for development of partitioned survival models in R. Results showed high accuracy with few or no errors but human intervention was needed.<br>Model adaptation: 1 study adapted an existing MS-Excel model developed for the UK setting to Czech Republic with overall accuracy of 97%. The update of the inputs were implemented with 100% accuracy. |
|  |  |  |  |  |  |  |  | Early HEE assessment: 1 study used GPT-4 in early modelling and reported effective processing of diverse data sources but without presenting clear results.<br>Optimisation and reporting: 1 study explored use of Copilot for MS-Excel to assist in developing VBA code within a health economic model. CoPilot was reported to successfully generate code and basic VBA tasks but was unable to meet more complex requirements specific to HEE. |
| O'Grady | 2024 | To identify the types of prompting techniques used in health economic modelling | Narrative review | Conference poster | Only studies that explicitly examined prompt engineering techniques were included | arXiv database | 13 | Three reviews summarised over 30 unique prompting techniques. Ten additional studies more closely examined 12 of these methods. Few-shot and chain-of-thought (COT) were shown to enhance response quality compared to zero-shot prompting. No studies were found where prompts were studied in an economic modelling context. |

#### Quality appraisal

The inadequate level of reporting inherent in conference abstracts and the absence of full publications precluded the use of the SANRA checklist for comprehensively appraising the included reviews. Common areas of inadequate reporting were in search strategy description, referencing and appropriate presentation of data. None of the reviews used more than 1 bibliographic database for the search and only 1 review (Dolin et al. 2024) used more than a single source, combining a database with conference and HTA web site sources.

#### Review synthesis

Boni et al. (2024) aimed to assess the uptake of AI in Health economic and outcome research (HEOR) based on studies presented at the ISPOR USA 2024 conference. A total of 17 (20%) of 83 studies identified were categorised as ‘AI-Driven Economic Modelling and Cost-Effectiveness’ with other categories more prevalent: ‘AI applications in health care’; ‘Data Extraction and Synthesis’ and in ‘Systematic Literature Reviews and Meta-Analyses’.

The review did not cite any included studies or report any further analysis on specific economic modelling use cases. It concluded that the findings showed a clear trend towards integrating AI within HEOR evidence generation and analysis across a wide range of applications.

Dolin et al. (2024) assessed how LLMs are currently applied in HEE modelling and HTA submissions. Of the 9 included case studies, 6 covered health economics applications, comprising model construction, model adaptation, model conceptualisation, input derivation and reporting of model results. All of these studies were assessed in our systematic review (see primary research) (Rawlinson 2024a, Rawlinson 2024b, Poirrier et al. 2023, Reason et al. 2024, Chhatwal et al. 2024a, Meissner et al. 2024) including one study identified through citation searching of this review (Meissner et al. 2024) which was ineligible. The results showed that reporting of methods and results was minimal. Performance, time or workload savings, and cost-effectiveness of LLM use were evaluated quantitatively in only 50% of HEE case studies and the code and data used were only reported by one study. The review did not report any results data from the included studies. Response accuracy was reported as the most common barrier to LLM implementation, with studies recommending the need for improving accuracy such as via prompt optimisation. Other barriers were data security, HTA body acceptance and the ease with which code produced by an LLM can be understood. The authors concluded that the use of LLMs for HEE and HTA is emerging, with minimal published evidence, which is exploratory and difficult to replicate.

A review (Fleurence et al. 2025), published as part of a working group report, introduced the uses of GenAI and foundation models, including LLMs, in the field of HTA. The studies relevant to economic modelling were all included in our review (Chatwal et al. 2024, Reason et al. 2024, Ayer et al. 2024), covering tasks of conceptualising, parameterising, construction and replicating studies. A key finding emerged that significant human supervision is needed in utilising LLMs for use in these tasks, to ensure accuracy and reliability.

Despite proof-of-concept research replicating a simple economic model, further research was recommended to fully replicate complex models. The review highlighted a further application of GenAI in the task of structural uncertainty analysis, with the potential for automating the process of addressing model weaknesses. The overall report strongly recommended that currently the use of GenAI tools should be restricted to supporting human activities with humans remaining accountable.

In a review of how GenAI has been applied in HEE, Lopez-Bernal et al. (2024) identified 4 of the studies (Reason et al. 2024, Rawlinson et al. 2024a, Srivastava et al. 2024b, Poirrier et al. 2023) included in our review. The use cases described were development, adaptation, early assessment, optimisation and reporting. The authors found that current application of GenAI is limited to improving existing methodological efficiency without extending to innovative modelling approaches. Key challenges to advance the field were reported as the need for robust validation methods, transparency, adaptation to diverse settings, and ethical considerations such as data privacy.

O’Grady et al. (2024) aimed to identify the types of prompting techniques used in HEE. Although over 30 prompting techniques were found from the 13 included studies, indicating a growing body of literature, no studies evaluated prompting techniques in the context of HEE. The authors recommended further research to address this evidence gap.

A broader review (Dasari et al. 2024) aimed to summarise the current use of AI and machine learning applications across different HEOR domains, with an emphasis on their impact on healthcare decision-making. Only 7 of the 105 included studies related to health economic modelling, the main applications comprising model automation, input validation, plot digitisation, code writing, and model conceptualisation. No further details were reported.

#### Expert opinion

One expert opinion piece (Srivastava et al. 2023) on the potential of LLMs in HTAs was identified. The abstract format precluded full critical appraisal, primarily due to inadequate reporting and lack of reference to, and discussion of, extant literature. LLM applications in HEE included conceptualising models by enabling testing of differing model structures, as well as optimising and validating models by iterative refinement. Challenges such as transparency, interpretability, bias and data quality were highlighted, in addition to ethical considerations including privacy protection and accountability.

## 4 Discussion

### Summary of findings

Our review has revealed that the current body of evidence on GenAI for HEE has been largely generated from recent conference paper submissions with minimal published findings from early case studies. The inadequate level of reporting inherent in conference abstracts precluded appraisal with the newly developed ELEVATE-AI LLMs framework (Fleurence et al. 2024) and highlights the still emergent nature of the evidence base. The included case studies, while contributing to early thinking on the topic, lack the detailed methodology, data, and analysis necessary for thorough evaluation, and the risk of bias through selective reporting cannot be ruled out.

The key outcomes of performance, time or workload savings, and error rates of LLM use were evaluated quantitatively in only 11 of the included case studies and the code and data used were only reported by the single fully published study. This was also the only study to report the time needed to rectify GenAI errors, a crucial factor that could offset efficiency savings of GenAI over a human-led approach. These reporting inadequacies, combined with the small number of case studies, prevented substantive conclusions on whether LLMs are appropriate for each of the use cases.

The absence of full peer-reviewed publications contrasts to the surge of recent case studies (94% in 2024) expedited as conference papers to disseminate results at specialist conferences. This highlights both the increasing topicality and anticipated impact of GenAI in the HEE context. The predominance of commercial authorship and industry funding of existing studies further indicates an emerging competitive market, with early manoeuvring of companies seeking to promote proof-of-concept products. An increasing number and range of GenAI tools could emerge in the coming years, accelerating the need for rigorous benchmarking and guidance for HTA agencies to select tools appropriately.

Replication, reported in 5 studies, was the most common use case, as might be expected at this early research stage without an established evidence base. Replication is effectively a transitory, gateway use case to validate the use of LLMs in HEE, before exploring their other capabilities. Despite early promising results, with low error rates, further replication studies will be needed to reinforce these findings, preferably in more diverse disease areas, and using different modelling approaches. They should also be designed to test LLMs in other use cases with higher impact in HEE. Questions also arose from these studies on the generalisability of results from the replication process to other types of economic models, including more complex models than the partitioned survival and Markov models predominantly explored in existing studies. Research was particularly sparse in the validation use case, where no studies were identified, thereby highlighting a research gap in this area.

While there were limited studies on the construction, updating and adaptation use cases, the reported results showed potential. This was especially evident in the 2 studies looking at using LLMs to adapt economic evaluations from one country to another, where high levels of accuracy were shown, even without the need for human intervention. However, the small number of studies in this area further underscores the need for additional confirmatory research to validate this use case. Evidence on the novel use case of data extraction from final appraisal documents in the post-submission phase, to inform market access analysis, was limited to 1 study. In this case the LLM failed to extract a substantial proportion of the necessary economic data from an appraisal report and further research is needed to explore these deficiencies.

Several studies explored optimising existing Excel based economic models using LLM powered plugins and interface assistance. The use of these plugins can conceivably help improve efficiency (by streamlining model code, thereby reducing runtime) and user experience, as well as reducing errors when updating existing economic models. However, current evidence is limited in the absence of quantitative outcome data.

Reporting and dissemination of results from an economic evaluation using LLMs were also explored in 2 studies. However, there was a lack of clarity on the prompt mechanisms used to perform these tasks, and accuracy of the end results, with no quantifiable accuracy metrics reported. Full publication of these studies may help to improve transparency and reproducibility.

### Implications for HTA practice

Studies have been exploratory with little potential to support the use of GenAI in HEE in HTA practice. GenAI methods remain at an early stage of maturity, with the tools yet to show demonstrable benefits in HTA decision-making. The current cluster of conference publications has served to disseminate early findings and stimulate discussion, but without fully transparent and reproducible reporting of coding and prompt mechanisms, this preliminary evidence will be of limited impact in practice to support the use of GenAI in HTA submissions. Furthermore, since most readers were not present at the conference sessions where the abstracts were presented, they would lack the additional context provided at the event to inform implementation.

Certain conference abstracts included at the title and abstract review stage were eventually excluded from our review based on inadequate reporting of health economic data in the abstract. This included studies referring to HTAs in the abstract, without clarity on which aspects of HTA they were focusing on, and studies exploring modelling techniques without specifying if health economic outcomes were reported. Had the studies been published in full, a more complete assessment of relevance and quality could be made.

The need to publish full results with prompt mechanisms is paramount if GenAI tools are to become embedded in routine economic evaluation practice. The secondary evidence in our review further reinforced this call for more transparent and reproducible reporting, with important research recommendations to address the gaps in evidence across the range of HEE use cases and prompting mechanisms. The need for standardised guidance in evaluating quality, transparency and rigour is being partly addressed by the development of ELEVATE-AI LLMs (Fleurence et al. 2024), the first framework dedicated to improving reporting standards of LLM-assisted research in this context for authors and reviewers. It builds on the foundational principles of existing frameworks for AI in healthcare, including HELM (Liang et al. 2022) and PALISADE (Padula et al. 2022), as well as emerging AI-specific guidelines such as CHEERS-AI (Elvidge et al. 2024), PRISMA-AI (Cacciamani et al. 2023), TRIPOD+AI (Collins et al. 2024) and RAISE (Thomas et al. 2024). Additionally, it complements international consensus guidelines, such as FUTURE-AI (Lekadir et al. 2025), to ensure rigorous standards for AI in healthcare.

Although copyright and licensing issues were not explored substantively in the primary and secondary studies, they remain ongoing barriers to implementation which must be addressed by researchers and regulatory bodies. Central to this is the need to establish and adhere to clear licensing agreements for the use of third-party content in training GenAI tools, as well as for accessing the GenAI code and prompt mechanisms used in evidence submissions. The NICE AI position statement (NICE, 2024a) sets out the responsibilities of submitting organisations in this area.

The need for human oversight was a prominent theme in our review with 6 of the included case studies either using a human-in-the-loop to cross check the performance of the LLM, or emphasising the need for it. This was reinforced by the included reviews and expert opinion which, in line with NICE’s position statement (NICE, 2024a), highlight the need for human expertise and LLMs to work in conjunction to produce high quality outputs.

### Areas for future research

The findings of our review indicate the pressing need for researchers to publish results in full where possible to advance knowledge in this expanding field. The paucity of fully published results and key gaps in the evidence base should serve to stimulate both new research and the translation of existing conference abstracts into full publications.

Research examining efficient implementation of human-in-the-loop approaches should be explored, aligning with the NICE position statement (NICE, 2024a) to maintain quality and trust in AI applications by augmenting rather than replacing human involvement. A key theme emerging from this review was the reported need for human interaction to complete HEE tasks. Future research should include the quantifiable reporting of the extent of human intervention, including time to rectify errors, to capture net time saved or incurred by using GenAI.

Further research is urgently needed in all use cases with testing in both exploratory and implementation phases. Replication studies should be followed up with application in key stages of modelling, particularly model construction and updating. With no studies on validation, the value of GenAI in this use case remains uncertain and represents a research gap. Ongoing <u>collaborative work</u> by NICE and industry partners will contribute to addressing this.

Additional unexplored use cases have also emerged in other HEE tasks which would not be viable without automation, including efficiently developing multiple model types to characterise a decision problem to quantify structural uncertainty (Fleurence et al. 2025); survival analysis involving extrapolation of time-to-event curves (Wu et al. 2024); and generation of synthetic data that replicates real patient data without personal identifiers, thereby protecting data privacy (Mosquera et al. 2023). Such novel applications may be part of the solution to overcoming existing barriers and GenAI itself may progressively adapt to address many of its own challenges.

However, there remains an urgent need to examine issues related to explainability, a particular challenge due to the ‘black box’ nature of LLMs often preventing a clear understanding by humans, which in turn leads to barriers of acceptance and ethical deployment (Marey et al. 2024). Further research is needed in the HEE context to explore prompt engineering to improve explainability of how LLMs generate outputs and ensure transparency of LLM decision making processes.

Accuracy and reliability concerns must be addressed through further testing and refinement of LLMs beyond proof-of-concept stages. This will include the use of feedback loops to continuously improve and fine tune model outputs, prompt optimisation through refining input data, and enhancing reliability by using techniques such as self-consistency (Taubenfeld et al. 2025).

Reproducibility and transferability is a further priority research area, with the need to explore how LLM prompts can be generalised and adapted across different decision problems and model types. Most of the existing studies have focused on either Markov or partitioned survival models and, while it is logical to first explore the capabilities of GenAI in these simpler modelling techniques, the more complex approaches, such as discretely integrated condition event models, remain unexplored. Evidence is needed to demonstrate the ability of LLMs to accommodate these diverse model types and to extend to a broader range of disease areas and scenarios reflective of HTA agency scope.

### Limitations

Our search was limited to published evidence. Application and testing of GenAI tools within the commercial sector may remain unpublished for competitive reasons. Reluctance to publish code and prompt mechanisms may remain a barrier to transparent publishing as companies prioritise protection of proprietary information and commercial interests.

The vast majority of included studies were submitted in conference abstract form, thereby precluding full quality assessment. Future iterations of the review will seek to apply the ELEVATE-AI LLMs framework for more comprehensive assessment of reporting in full publications.

Our review protocol did not include contacting authors for further information on their publications. Authors are expected to publish their complete results whenever possible, and to extend beyond the limited dissemination of conference abstracts. As such, it is anticipated that the studies included in our review will translate into full publications over time, for more complete assessment in our future update.

Despite these limitations, our review has several strengths. To our knowledge it is the first systematic literature review on this topic and sets out the early evidence landscape at the time of publication to provide a foundation for future research. By highlighting evidence gaps, the review recommendations aim to stimulate further meaningful research in priority areas and thereby encourage the development of the evidence ecosystem on the topic. This will be further enhanced by our planned living approach to updating the review over time to capture ongoing developments. The inclusive scope of the review, extending beyond economic modelling to HEE, has also allowed exploration of potential novel use cases of GenAI that were previously unfeasible under a standard approach. Finally, incorporating both primary and secondary evidence to draw upon the totality of available data has identified several key barriers and challenges to be addressed before GenAI can be implemented in practice.

## 5 Conclusion

In the nascent but rapidly developing field of GenAI in HEE, this review has established the current evidence landscape, revealing potential value across multiple use cases but with only limited reporting of early case studies via conference submissions. Research to date has focused on exploratory studies to validate GenAI tools without fully demonstrating their added value in key stages of modelling. As such, drawing definitive conclusions would be premature and further confirmatory evidence is awaited across all use cases. Key barriers and challenges must be addressed through peer-reviewed research and international collaboration to develop best practice recommendations and standards. In this dynamic environment, further updates to the review will capture emerging thinking and innovation to inform guidance for HTA agencies to navigate the increasing range and complexity of GenAI tools.

## Glossary of AI and health economic terms

- Model adaptation: Modifying the model for different jurisdictions, settings, populations, and treatment pathways.
- Artificial intelligence (AI): A broad field of computer science that aims to create intelligent machines capable of performing tasks typically requiring human intelligence.
- Chain of thought prompting: A technique where the model is guided to reason through a problem step-by-step in its response, by breaking down complex tasks into simpler parts to improve accuracy.
- Model conceptualisation: Designing the model structure including type of model, health states to be included and transitions between health states (where applicable) ensuring comprehensive coverage of relevant factors and scenarios.
- Deep Learning: A subset of machine learning algorithms that uses multilayered neural networks, called deep neural networks. Those algorithms are the core behind the majority of advanced AI models.
- Model construction: Creating the structure and framework of the health economic model, including defining the model’s components, such as states, transitions, and time horizons, and implementing the model using appropriate software and algorithms.
- Few-Shot Learning: A method where a model is given a few examples in the prompt to guide its response, utilizing its pre-trained knowledge to produce accurate outputs with minimal data.
- Foundation model: A large scale pretrained models that serve a variety of purposes. These models are trained on broad data at scale and can adapt to a wide range of tasks and domains with further fine-tuning.
- Generative AI: AI systems capable of generating text, images, or other content based on response to user prompts. It uses machine learning techniques to create new data that has similar characteristics to the data it was trained on.
- Generative Pre-trained Transformer (GPT): A specific series of LLMs created by OpenAI based on the Transformer architecture, which is particularly well-suited for generating human-like text.
- Hallucination: Large language models, such as ChatGPT, are unable to identify if the phrases they generate make sense or are accurate. This can sometimes lead to inaccurate results, also known as ‘hallucination’ effects, where large language models generate plausible sounding but inaccurate text. Hallucinations can also result from biases in training datasets or the model’s lack of access to up-to-date information
- Incremental cost-effectiveness ratio (ICER): The incremental cost-effectiveness ratio (ICER) is the difference in the change in mean costs in the population of interest divided by the difference in the change in mean outcomes in the population of interest.
- Large Language Model: A specific type of foundation model trained on massive text data that can recognise, summarise, translate, predict, and generate text and other content based on knowledge gained from massive datasets.
- Markov modelling: A decision-analytic technique that characterises the prognosis of a group by assigning group members to a fixed number of health states and then modelling transitions among the health states.
- Machine learning (ML): A field of study within AI that focuses on developing algorithms that can learn from data without being explicitly programmed.
- Multimodal AI: An AI model that simultaneously integrates diverse data formats provided as training and prompt inputs, including images, text, bio-signals, -omics data and more.
- Model optimisation: Adjusting the model parameters, formulae, and decision-making criteria to achieve the most efficient and accurate outcomes. This involves refining inputs and improving model structure or assumptions.
- Prompt: The input given to an AI system, consisting of text or parameters that guide the AI to generate text, images, or other outputs in response.
- Prompt engineering: Creating and adapting prompts (input) to instruct AI models to generate specific output.
- Quality-adjusted life year (QALY): A measure of the state of health of a person or group in which the benefits, in terms of length of life, are adjusted to reflect the quality of life. One quality-adjusted life year (QALY) is equal to 1 year of life in perfect health. QALYs are calculated by estimating the years of life remaining for a patient following a particular treatment or intervention and weighting each year with a quality-of-life score (on a 0 to 1 scale). It is often measured in terms of the person’s ability to carry out the activities of daily life, and freedom from pain and mental disturbance.
- Model replication: Generating a replica of a published model to facilitate other use cases, including development, adaptation, validation, and adaptation.
- Model reporting: Providing interpretations, contextualising results, generating reports and visualisations for stakeholders.
- Retrieval Augmented Generation (RAG): A method in natural language processing that combines a generative model with a retrieval system to improve response accuracy. The retrieval system finds relevant information, which the generative model uses to produce more contextually accurate and factual outputs.
- Sensitivity Analysis: A means of exploring uncertainty in the results of economic evaluations. There may be uncertainty because data are missing, estimates are imprecise or there is controversy about methodology. Sensitivity analysis can also be used to see how applicable results are to other settings. The analysis is repeated using different assumptions to examine the effect of these assumptions on the results.
- Model updating: Updating a model with new data and evidence, ensuring that the models remain current and accurate over time.
- Model validation: Assessing whether the model accurately represents the real-world system it is intended to simulate. Internal validation involves quality assessing the model against prespecified criteria. External validation compares the model’s outputs with external data not used in the model’s development. Sources: Fleurence et al. 2025, Parliamentary Office of Science and Technology, NICE Glossary

## Appendix A: Search strategy

The full search strategy for Medline ALL (Ovid platform) is given below. This was adapted for use in other databases, taking into account variations in index terms (where available). Search strategies for all databases (Embase – Ovid platform; Econlit – Ovid platform; the Cochrane Database of Systematic Reviews – Wiley platform; INAHTA International HTA database; Epistemonikos; Europe PMC [for preprints only] and PROSPERO) are available on request. Searches were last updated between the 3^rd^ and 5^th^ March 2025.

### Ovid MEDLINE(R) ALL <1946 to February 28, 2025>

1. exp *models, economic/ 5494
2. ((economic or pharmacoeconomic or “cost-benefit” or “cost comparison” or “cost comparisons” or “cost-effectiveness” or “cost-minimisation” or “cost-minimization” or “cost-utility” or “cost-consequence” or “comparative effectiveness”) adj (model or models or modelling or analysis or analyses or evaluation or evaluations or research or study or studies)).ti,ab,kf.63546
3. *Cost-Effectiveness Analysis/ 283
4. *Cost-Benefit Analysis/mt 1554
5. *“Costs and Cost Analysis”/mt 550
6. exp *Technology Assessment, Biomedical/mt 1132
7. “health technology assessment”.ti,ab,kf. 7376
8. “health technology assessments”.ti,ab,kf. 746
9. “health technology evaluation”.ti,ab,kf. 41
10. “health technology evaluations”.ti,ab,kf. 4
11. “technology appraisal”.ti,ab,kf. 471
12. “technology appraisals”.ti,ab,kf. 160
13. HTA.ti,ab,kf. 4727
14. HTAs.ti,ab,kf. 483
15. *Comparative Effectiveness Research/mt 454
16. or/1-15 76285
17. exp *Artificial Intelligence/ 139957
18. “generative artificial”.ti,ab,kf. 931
19. “generative AI”.ti,ab,kf. 1079
20. “AI-generated”.ti,ab,kf. 748
21. “gen-AI”.ti,ab,kf. 14
22. genAI.ti,ab,kf. 106
23. GAI.ti,ab,kf. 816
24. “large language model”.ti,ab,kf. 1726
25. “large language models”.ti,ab,kf. 3786
26. LLM.ti,ab,kf. 1943
27. LLMs.ti,ab,kf.2177
28. “foundation model”.ti,ab,kf. 292
29. “foundation models”.ti,ab,kf. 308
30. “transformer-based model”.ti,ab,kf. 228
31. “transformer-based models”.ti,ab,kf. 225
32. “retrieval-augmented generation”.ti,ab,kf. 146
33. “neural net”.ti,ab,kf. 643
34. “neural nets”.ti,ab,kf. 465
35. “neural network”.ti,ab,kf. 88391
36. “neural networks”.ti,ab,kf. 61017
37. “machine learning”.ti,ab,kf. 140424
38. “deep learning”.ti,ab,kf. 80323
39. “unsupervised learning”.ti,ab,kf. 3380
40. “unsupervized learning”.ti,ab,kf. 1
41. “generative adversarial network”.ti,ab,kf. 3098
42. “generative adversarial networks”.ti,ab,kf. 2282
43. gan.ti,ab,kf. 9496
44. gans.ti,ab,kf. 1366
45. “generative pretrained transformer”.ti,ab,kf. 249
46. “generative pretrained transformers”.ti,ab,kf. 32
47. “generative pre-trained transformer”.ti,ab,kf. 612
48. “generative pre-trained transformers”.ti,ab,kf. 68
49. gpt*.ti,ab,kf. 8091
50. ChatGPT*.ti,ab,kf. 5698
51. Chat-GPT*.ti,ab,kf. 177
52. “variational autoencoder”.ti,ab,kf. 995
53. “variational autoencoders”.ti,ab,kf. 395
54. vae.ti,ab,kf. 1251
55. vaes.ti,ab,kf. 353
56. “Bi-directional encoder representations from transformers”.ti,ab,kf. 4
57. “Bidirectional encoder representations from transformers”.ti,ab,kf. 656
58. BERT.ti,ab,kf. 1928
59. “multimodal model”.ti,ab,kf. 240
60. “multimodal models”.ti,ab,kf. 138
61. “multi-modal model”.ti,ab,kf. 55
62. “multi-modal models”.ti,ab,kf. 30
63. “pathways language model”.ti,ab,kf. 4
64. (Google and palm*).ti,ab,kf. 113
65. medpalm*.ti,ab,kf. 1
66. “med-palm”.ti,ab,kf. 14
67. “med-palm-2”.ti,ab,kf. 5
68. gemini*.ti,ab,kf. 5540
69. bard*.ti,ab,kf. 6242
70. llama*.ti,ab,kf. 2701
71. claude*.ti,ab,kf. 1672
72. falcon*.ti,ab,kf. 1949
73. mixtral*.ti,ab,kf. 26
74. chatbot*.ti,ab,kf. 2692
75. “virtual assistant”.ti,ab,kf. 163
76. “virtual assistants”.ti,ab,kf. 139
77. “virtual agent”.ti,ab,kf. 170
78. “virtual agents”.ti,ab,kf. 194
79. or/17-78 387512
80. 16 and 79 574
81. animals/ 7610901
82. exp Animals, Laboratory/ 989323
83. exp Animal Experimentation/ 10666
84. exp Models, Animal/ 674985
85. exp Rodentia/ 3688804
86. (rat or rats or mouse or mice or rodent*).ti. 1522283
87. or/81-86 7742424
88. 87 not humans/ 5399792
89. 80 not 88 567
90. limit 89 to english language 557
91. limit 90 to letter 4
92. 90 not 91 553

## Appendix B: PRISMA study selection flow chart

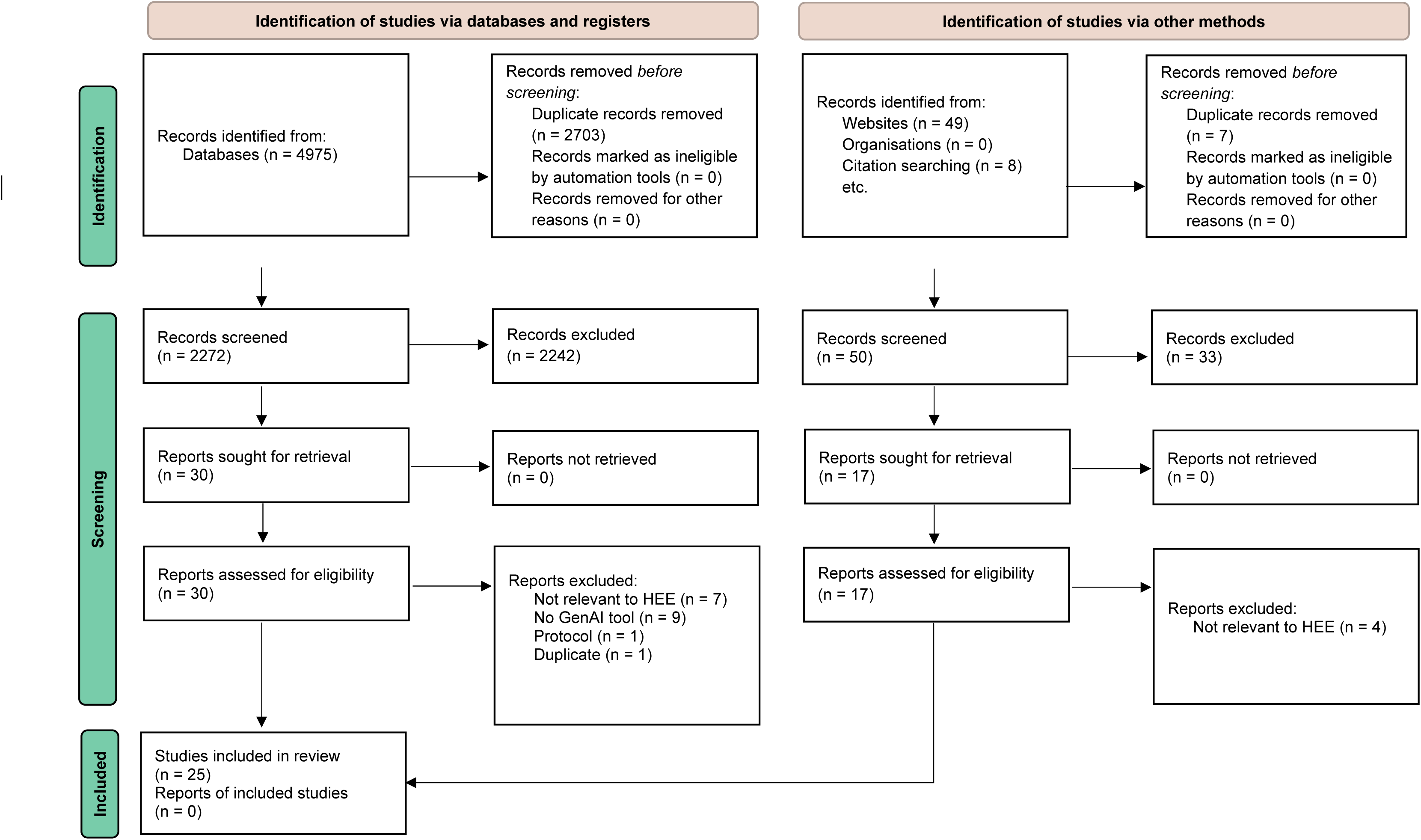

## Data Availability

All data produced in the present work are contained in the manuscript

